# Retrospective Analysis of an Acute Ischemic Stroke Cohort Shows that Timing of Antidepressant Use Associated with Short-term Recovery and Functional Independence at 90-Days

**DOI:** 10.1101/2024.02.08.24302550

**Authors:** Elizabeth Baraban, Alexandra Lesko, Kyle Still, Weston Anderson

## Abstract

**Background:** Little is known about how the timing of antidepressant use influences stroke outcomes. The objective of this exploratory, retrospective analysis is to examine stroke outcomes by timing of antidepressant use among patients who received stroke treatment.

**Methods:** 12,590 eligible patients were treated for a primary or secondary diagnosis of ischemic stroke. Inpatient strokes and patients who were missing information on antidepressant use or stroke outcomes were excluded. The outcome variables were (1) a change in ambulation from pre-stroke to discharge; (2) a change in modified Rankin scale (mRS) from pre-stroke to discharge; and (3) a change in mRS from pre-stroke to 90-days post-discharge. The independent variable of interest was timing of antidepressant treatment. Logistic regression with generalized estimating equations was used, controlling for covariates.

**Results:** At 90-days post-discharge, patients with no history of antidepressant use or with a previous history of antidepressant use were similar to patients with current antidepressant use in terms of their return to baseline functional independence. In contrast, our model predicted that a new antidepressant prescription at discharge was associated with a ∼57% decrease in the likelihood of returning to baseline functional independence at 90-days compared to patients currently using an antidepressant (AOR:0.510, CI:0.277-0.938, p=0.03). Post-hoc analyses showed females with current antidepressant use associated with a higher predicted probability of returning to baseline functional independence at 90-days compared to females with no antidepressant use. This association was not true for males. Conversely, males with a new prescription had the lowest predicted probability of returning to baseline among all groups at 90-days post-discharge.

**Conclusion:** These results suggest that use of antidepressants was associated with stroke recovery, but the effects are moderated by sex. Further study is needed to determine if this relationship is causal and the mechanisms between timing of antidepressant treatment and outcomes.

## 1.0 Introduction

Published research suggests that antidepressants may improve outcomes in survivors of acute ischemic stroke. Specifically, the FLAME study found that post-stroke treatment with fluoxetine combined with psychical therapy enhanced motor recovery after three months compared to those treated with placebo.[1] Also, a 2012 meta-analysis found that antidepressant use post-stroke was associated with greater independence, more functionality, less neurological impairment, less anxiety, and less depression.[2] A proposed explanation for this relationship is that antidepressants may increase neuro-regeneration. In support of this idea, a meta-analysis to determine the efficacy of antidepressants in animal models of ischemia showed that antidepressants improved infarct volume, neurobehavioral outcomes, and neurogenesis.[3] Another theory is that antidepressant use improves patients’ mood and motivation to perform rehabilitative tasks. This is thought to help facilitate recovery.[4]

However, more recent studies revealed conflicting results. The AFFINITY (2020), FOCUS (2019), and EFFECTS (2020) trials found that fluoxetine for six months daily after a stroke does not improve functional outcomes. [5–7] Furthermore, an updated meta-analysis in 2019 looked at 63 different eligible trials exploring antidepressant use for stroke recovery. The authors found no reliable evidence that antidepressants should be used to promote recovery after stroke. [8]

To date, most studies, including those summarized above, focused on antidepressant use post-stroke. Far less is known about how the timing of antidepressant use associates with stroke outcomes. However, a few published studies suggest that antidepressant use prior to stroke may negatively impact outcomes. A 2010 study in patients with acute ischemic stroke treated with tPA found that prior antidepressant use associated with less favorable outcomes.[4] Another study found that antidepressant use pre-stroke is associated with an increased risk of a more severe stroke and death within 30 days.[9] In contrast, a 2015 study found antidepressant use before a stroke had a positive impact on stroke outcomes. Specifically, the authors found that patients who had antidepressant treatment prior to their stroke (n = 51) had better functional outcomes at discharge compared to those prescribed antidepressant treatment during their hospitalization (n = 188).[10]

Given that an estimated 13.2% of adults in the U.S. have used antidepressants, it is important to better understand how timing of antidepressant use associates with stroke outcomes.[11] The objective of this retrospective analysis is to examine post-stroke ambulation and functional independence by timing of antidepressant use among patients who received treatment for an acute ischemic stroke.

## 2.0 Materials and Methods

### 2.1 Data Source and Study Population

Data from hospitals in a multi-state healthcare system including Washington, Oregon, California, Texas, Montana, and Alaska were used. Diagnostic and outcome patient data were manually abstracted from each hospital’s medical record and entered in a system-wide stroke registry.

Eligible subjects were IV-thrombolytic or mechanical thrombectomy treated patients presenting with a primary or secondary diagnosis of acute ischemic stroke. For inclusion in the study, these patients were required to be over the age of 18 and discharged from the hospital between January 1, 2014, through December 31, 2021. Inpatient strokes and patients who were missing information on antidepressant use or stroke outcomes were excluded (Figure 1).

**Figure 1.**
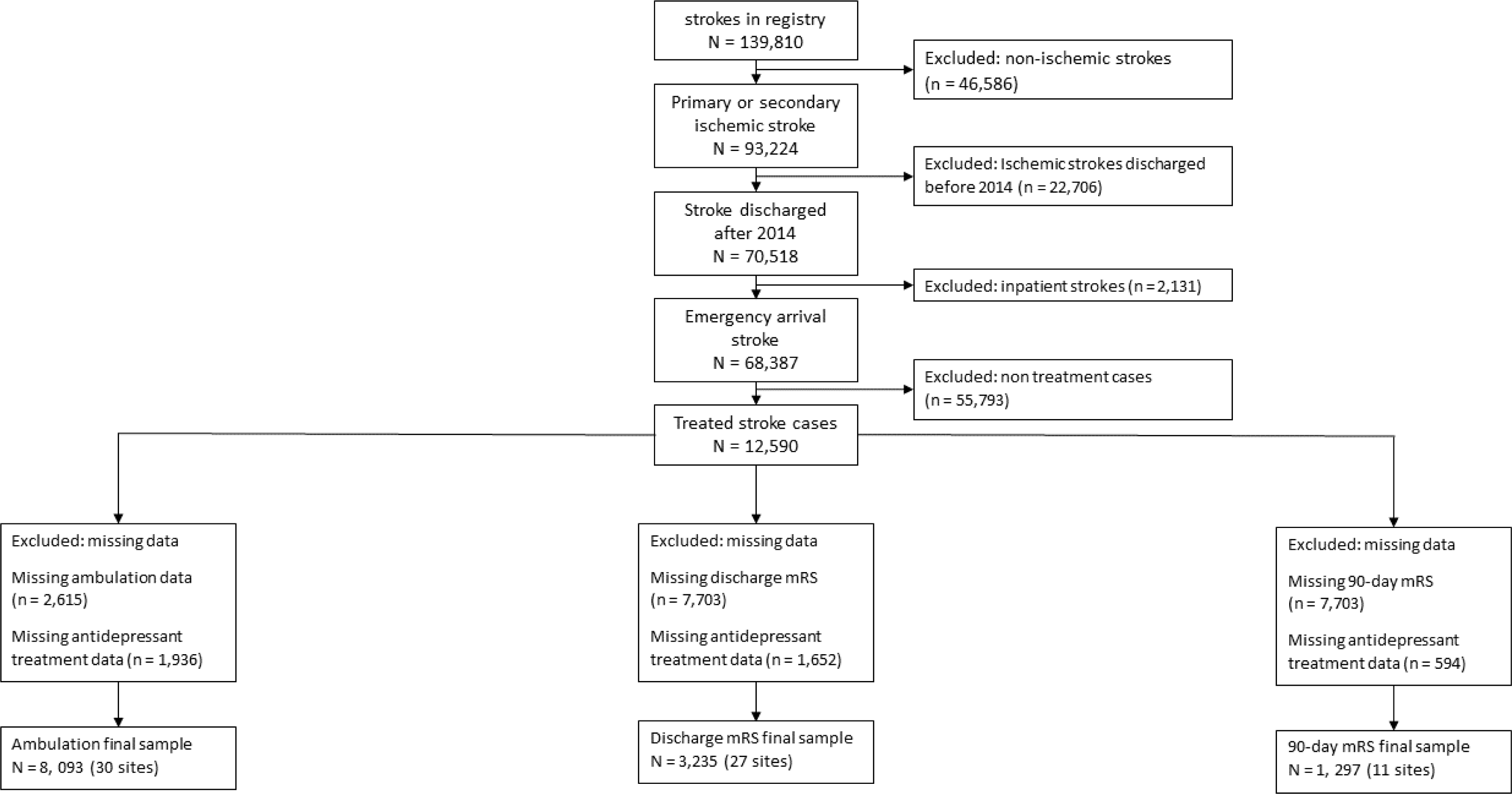
Cohort diagram: This flowchart shows how our exclusion criteria were applied to result in our three analytic samples for ambulation, discharge mRS, and 90-day mRS.

### 2.2 Variables

The outcome variables were:

- A change in ambulation from pre-stroke to discharge.
- A change in functional independence from pre-stroke occurrence to discharge.
- A change in functional independence from pre-stroke occurrence to 90-days after discharge.

Ambulation was determined using a three-item scale (able to ambulate independently, able to ambulate with help from another person, unable to ambulate) at pre-stroke and discharge.

Change in ambulation was dichotomized and categorized as “yes, returned to pre-stoke ambulation (baseline)” or “no, did not return to baseline ambulation.” Functional independence was measured using a dichotomized modified Rankin scale (mRS) for pre-stroke, at discharge, and at 90-days post-discharge. Patients with a mRS score of 0-2 were categorized as “able to look after themselves without assistance” and patients with a score of 3-5 were categorized as “needed assistance for daily living activities”. Change in mRS was dichotomized as “returned to baseline functional independence at 90-days” or “did not return to baseline functional independence at 90-days”.

The independent variable of interest was use of any class of antidepressant medication, which we categorized in five groups based on timing of use (Figure 2):

1. Prior antidepressant use only, defined as a history of antidepressant use but no documentation of antidepressant use within the week before stroke occurrence.
2. Current antidepressant use, defined as documented antidepressant use within the week before stroke occurrence and prior to hospital arrival.
3. New prescription at discharge, defined as discharge from stroke admission with a new antidepressant prescription but without a prescribed antidepressant at the time of stroke.
4. No antidepressant use, defined as a lack of documented antidepressant use at any time in the patient’s medical record.
5. No current antidepressant use, defined as a combination of groups 1 and 4.

**Figure 2.**
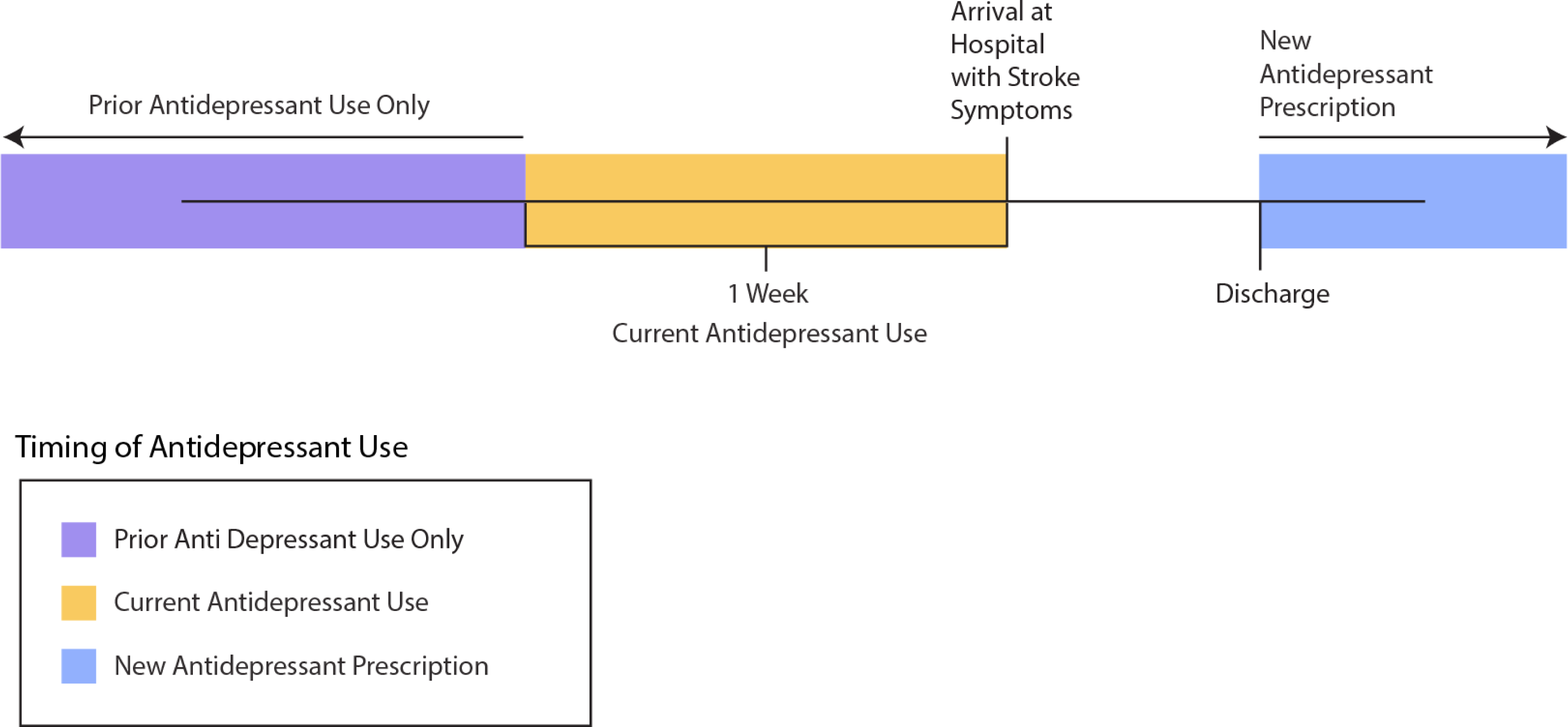
Timeline of Antidepressant Use: This figure shows a visual timeline of antidepressant use. Prior antidepressant use only was defined as a history of antidepressant use but no documentation of antidepressant use within the week before stroke occurrence. Current antidepressant use was defined as documented antidepressant use within the week before stroke occurrence and prior to hospital arrival. A new prescription at discharge was defined as discharge from stroke admission with a new antidepressant prescription but without a prescribed antidepressant at the time of stroke.

Antidepressant use and timing were abstracted from electronic medical records at arrival and discharge. Antidepressant medication classes included tricyclic and tetracyclic antidepressants, selective serotonin reuptake inhibitors, monoamine oxidase inhibitors, norepinephrine reuptake inhibitors, norepinephrine-dopamine reuptake inhibitors serotonin receptor antagonists/agonists, and 2-adrenergic receptor antagonists.

### 2.3 Statistical Analyses

Patient characteristics for change in ambulation and functional independence were summarized as percentages and medians [IQR]. Comparisons were made using chi-square or Fisher’s exact tests and t-tests or Mann-Whitney tests, as appropriate. Logistic regression with generalized estimating equations was used to identify the association of antidepressant use on outcomes. We controlled for covariates, which were chosen based on literature review and bivariate analysis. Covariates included age, sex, race and Hispanic ethnicity, National Institute of Health Stroke Scale (NIHSS) at arrival, time from last known well to hospital arrival, type of treatment (IV alteplase only versus mechanical thrombectomy only or both), insurance (private versus any Medicare versus any Medicaid versus self-pay/no insurance/unable to determine) and any medical history of diabetes, atrial fibrillation, smoking or obesity categorized as yes/no. Race and ethnicity were dichotomized as “White, non-Hispanic” and “racial and ethnic minorities,” which included American Indian or Native American, Asian, Black, Hispanic or Latino, and Native Hawaiian or Pacific Islander. For nonlinear, continuous covariates, polynomial terms were tested and added as appropriate.

### 2.4 Post-hoc Analyses

Post-hoc analyses were conducted to further investigate the relationship between antidepressant treatment and stroke outcomes. To assess whether the association between antidepressant use and change in ambulation or functional independence was influenced by a second predictor, we tested for interactions between antidepressant use and all other significant predictors one at a time. All variables that were included in the main effects model were also added as covariates to these models and adjustments for multiple comparisons were made. Predicated probabilities of returning to baseline status are reported.

To verify the appropriateness of the models, diagnostics of the model were examined. During model building, all tests were 2-tailed with alpha equal to .05; p-values < 0.05 were considered statistically significant. SPSS v26.0 software (SPSS Inc., Chicago, IL) were used for statistical analyses and graphics.

## 3.0 Results

### 3.1 Demographic data

From January 1, 2014, through December 31, 2021, 12,590 patients from 30 hospitals in the Providence St. Joseph Health network received treatment for a primary or secondary diagnosis of acute ischemic stroke. Figure 1 illustrates how we arrived at a final analytic sample for our outcome variables. Our total stroke population was 49.6% female, 21.5% were racial and ethnic minorities, and 75.5% had a medical history of one or more of the following conditions: atrial fibrillation, smoking, diabetes, or obesity (Table 1). The demographic and clinical characteristics of our outcomes-based analytic samples and the total stroke population were similar (Table 1). Current antidepressant use was also similar between our analytic samples.

**Table 1:**
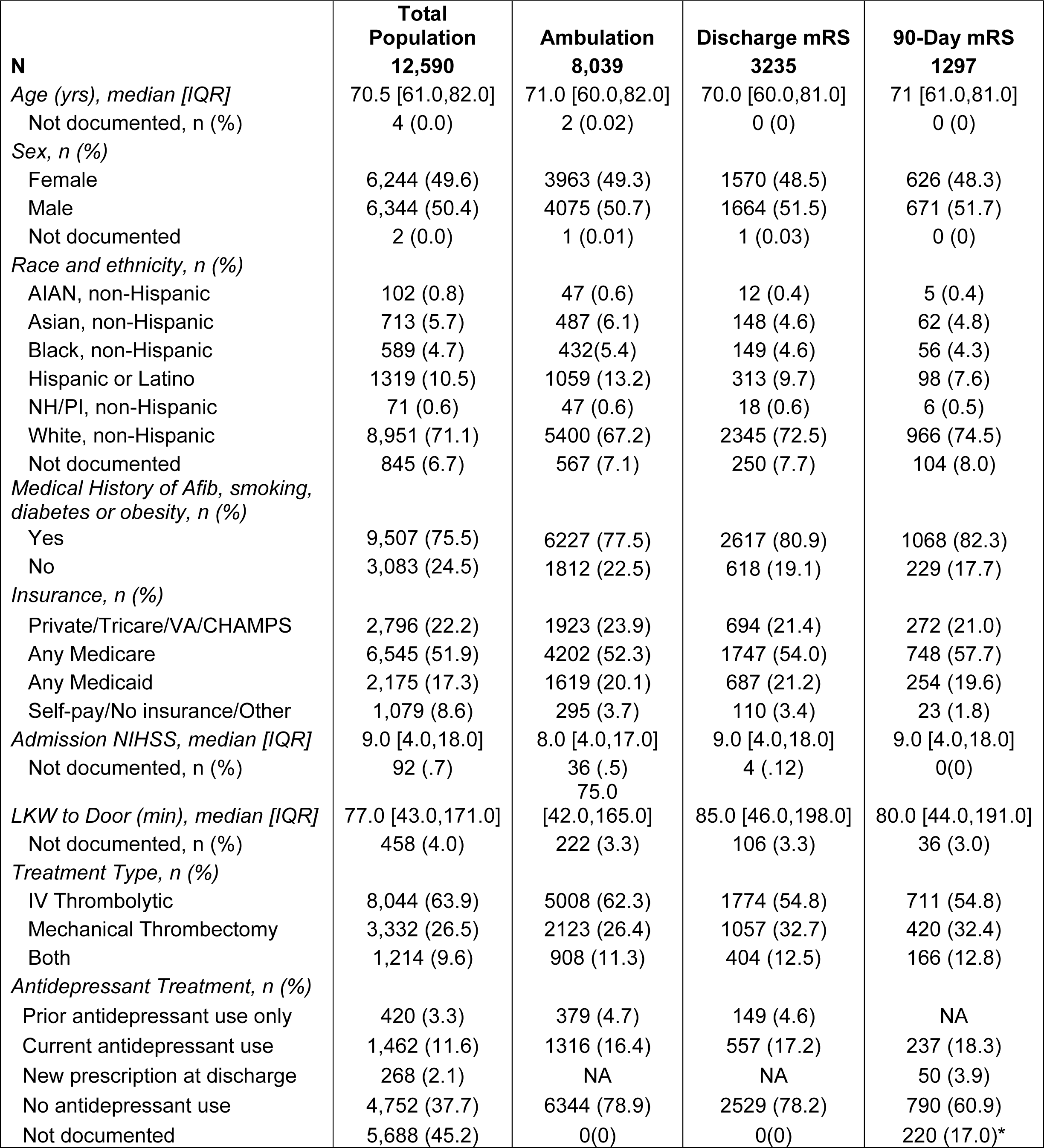
Patient characteristics at admission.

### 3.2 Change in ambulation from pre-stroke to discharge

The final study sample for ambulation included 8,039 patient encounters (Table 1). 54.8% (n = 4,403) returned to baseline ambulation at time of discharge (Table 2). Compared to patients who returned to baseline ambulation, patients with worse ambulation at discharge were older (67 vs 76, p = <.001), had a higher NIHSS score at arrival (6 vs 14, p = <.001), and had a longer last known well to arrival time (69 min vs 90min, p = <.001, Table 2). Notably, fewer patients with worse ambulation at discharge had current antidepressant use (p = 0.007, Table 2). Additionally, a greater proportion of patients with worse ambulation at discharge were female or racial and ethnic minorities. Multiple logistic regression analyses showed that patients with no current antidepressant use were associated with a decreased likelihood of returning to baseline ambulation at discharge compared to those who were currently on an antidepressant (AOR=0.795; 95% CI 0.679-0.930). Likewise, compared to patients currently on an antidepressant, those with prior antidepressant use only (AOR=0.641; 95% CI 0.461-0.893) or no antidepressant use (AOR=0.807; 95% CI 0.691-0.942) associated with a decreased likelihood of returning to baseline ambulation at discharge (Table 3).

**Table 2:** Cohort characteristics by ambulation and mRS at discharge.

|  | Change in Ambulation from Pre-Stroke to Discharge |  |  | Change in mRS from Pre-Stroke to Discharge |  |  |
| --- | --- | --- | --- | --- | --- | --- |
|  | Returned to Baseline | Worsened | p-value* | Returned to Baseline | Worsened | p-value* |
| **n (%)** | 4403 (54.8) | 3636 (45.2) |  | 1560 (48.2) | 1675 (51.8) |  |
| Age (yrs), median [IQR] | 67.0 [56.0,77.0] | 76.0 [65.0,85.0] | <.001 | 69.0 [58.0,79.0] | 72.0 [62.0,82.0] | <.001 |
| Sex, n (%) |  |  |  |  |  |  |
| Female | 2019 (50.9) | 1944 (49.1) | <.001 | 753 (48.0) | 817 (52.0) | 0.787 |
| Male | 2384 (58.5) | 1691 (41.5) |  | 806 (48.4) | 858 (51.6) |  |
| Race/ethnicity, n (%) |  |  |  |  |  |  |
| White, non-Hispanic | 3078 (57.0) | 2322 (43.0) | <.001 | 1161 (49.5) | 1184 (50.5) | 0.017 |
| Racial and Ethnic Minorities | 1325 (50.2) | 1314 (49.8) |  | 399 (44.8) | 491 (55.2) |  |
| Medical History of Afib, smoking, diabetes or obesity, n (%) |  |  |  |  |  |  |
| Yes | 3402 (54.6) | 2825 (45.4) | 0.646 | 1271 (48.6) | 1346 (51.4) | 0.42 |
| No | 1001 (55.2) | 811 (44.8) |  | 289 (46.8) | 329 (53.2) |  |
| Insurance, n (%) |  |  |  |  |  |  |
| Private/Tricare/VA/CHAMPS | 1244 (64.7) | 679 (35.3) | <.001 | 408 (59.0) | 283 (41.0) | 0.42 |
| Any Medicare | 2085 (49.6) | 2117 (50.4) |  | 768 (44.0) | 979 (56.0) |  |
| Any Medicaid | 881 (54.4) | 738 (45.6) |  | 328 (47.7) | 359 (52.3) |  |
| Self-pay/No insurance/Other | 193 (65.4) | 102 (34.6) |  | 56 (50.9) | 54 (49.1) |  |
| Admission NIHSS, median [IQR] | 6.0 [3.0,11.0] | 14.0 [7.0,22.0] | <.001 | 6.0 [3.0,12.0] | 13.0 [6.0,20.0] | <.001 |
| LKW to Door (min), median [IQR] | 69 [46.0,136.0] | 90 [46.0,231] | <.001 | 70 [41.0,141.0] | 120.0 [51.3,295.8] | <.001 |
| Treatment Type, n (%) |  |  |  |  |  |  |
| IV Thrombolytic | 3179 (63.5) | 1829 (36.5) | <.001 | 1076 (60.7) | 698 (39.3) | <.001 |
| Mechanical Thrombectomy | 788 (37.1) | 1335 (62.9) |  | 319 (30.2) | 738 (69.8) |  |
| Both | 436 (50.2) | 472 (48.0) |  | 165 (40.8) | 239 (59.2) |  |
| Antidepressant Use, n (%) |  |  |  |  |  |  |
| Current antidepressant use | 765 (58.1) | 551 (41.9) | 0.007 | 314 (56.4) | 243 (43.6) | <.001 |
| No current use | 3638 (54.1) | 3085 (45.9) |  | 1246 (46.5) | 1432 (53.5) |  |
| Current antidepressant use | 765 (58.1) | 551 (41.9) | 0.017 | 314 (56.4) | 243 (43.6) | <.001 |
| Prior antidepressant use only | 196 (51.7) | 183 (48.3) |  | 65 (43.6) | 84 (56.4) |  |
| No antidepressant use | 3442 (54.3) | 2902 (45.2) |  | 1181 (46.7) | 1348 (53.3) |  |

Racial and Ethnic Minorities: American Indian or American Native, Asian, Black, Hispanic, Native Hawaiian or Pacific Islander; Afib: atrial fibrillation; VA: Veterans Affairs; CHAMPS: Community Health Automated Medicaid Processing System; NIHSS: National Institutes of Health Stroke Scale; LKW: Last known well.

**Table 3:**
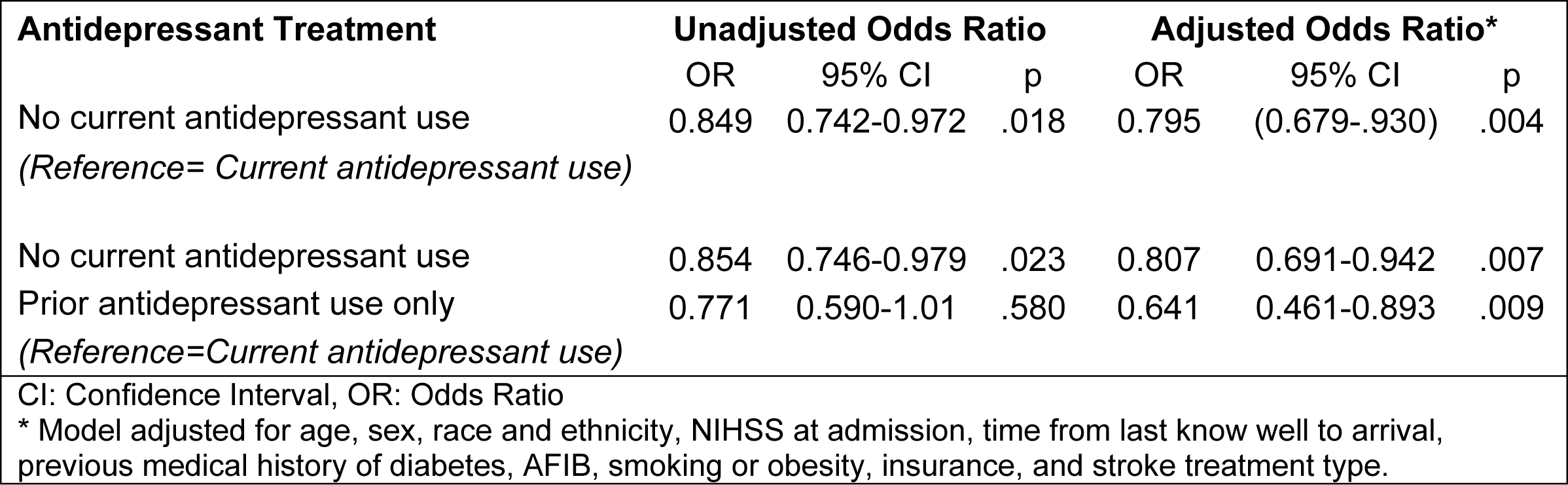
Model of change in ambulation (pre-stroke to discharge)

### 3.3 Change in functional independence from pre-stroke to discharge

The final study sample for modified Rankin at discharge included 3,235 encounters (Table 1). 51.8% (n = 1,675) had worse functional independence at discharge (Table 2). Compared to patients who returned to baseline function status, those with worse functional independence at discharge were older (69 vs 72, p = <.001), had worse NIHSS scores at arrival (6 vs 13, p = <.001), and had longer last known well to door times (70min vs 120min, p = <.001). Notably, fewer patients with worse functional independence at discharge were currently on an antidepressant (p = <.001, Table 2). Additionally, a greater proportion of patients who did worse were racial and ethnic minorities. The multivariate analyses showed that patients who had no current antidepressant were associated with a decreased likelihood of returning to baseline functional independence at discharge compared to those who were currently on an antidepressant (AOR= 0.681; 95% CI 0.488-0.949). Likewise, compared to patients currently on an antidepressant, those with prior antidepressant use only (AOR=0.693; 95% CI 0.494-0.971) or no antidepressant use (AOR=0.532; 95%CI 0.360-0.785) were associated with a decreased likelihood of returning to baseline discharge (Table 4).

**Table 4:** Model of change in mRS (pre-stroke to discharge)

| Antidepressant Treatment | Unadjusted Odds Ratio |  |  | Adjusted Odds Ratio* |  |  |
| --- | --- | --- | --- | --- | --- | --- |
|  | OR | 95% CI | p | OR | 95% CI | p |
| No current antidepressant use<br>*Reference=Current antidepressant use)* | 0.673 | 0.529-0.858 | .001 | 0.681 | 0.488-0.949 | .023 |
| No current antidepressant use | 0.678 | 0.532-0.865 | .002 | 0.693 | 0.494-0.971 | .033 |
| Prior antidepressant use only<br>*(Reference=Current antidepressant use)* | 0.599 | 0.437-0.823 | .001 | 0.532 | 0.360-0.785 | .001 |
| mRS: Modified Rankin Scale, CI: Confidence Interval, OR: Odds Ratio |  |  |  |  |  |  |
| * Model adjusted for age, sex, race and ethnicity, NIHSS at admission, time from last know well to arrival, previous medical history of diabetes, AFIB, smoking or obesity, insurance, and stroke treatment type. |  |  |  |  |  |  |

### 3.4 Change in functional independence from pre-stroke to 90-days post-discharge

The final study sample for 90-day modified Rankin included 1,297 encounters (Table 1); 65.0% (n=843) of these patients returned to baseline functional independence at 90-days post-discharge (Table 5). Compared to those that returned to baseline functional independence at 90-days, patients that did worse at 90-days were older (70 vs 75, p=<.001), had worse NIHSS scores at arrival (6 vs 16, p=<.001), and had longer last known well to door times (71min vs 118min, p=<.001, Table 5). Additionally, a greater proportion of patients that worsened were female and racial and ethnic minorities (Table 5). Multivariate analyses showed no association of current antidepressant use at the time of the stroke with 90-day functional independence (AOR=0.959; 95%CI 0.756-0.1.31). However, patients prescribed antidepressants at discharge were associated with a decreased likelihood of returning to baseline functional independence at 90-days (AOR=0.510; 95%CI 0.277-0.938, Table 6).

**Table 5:**
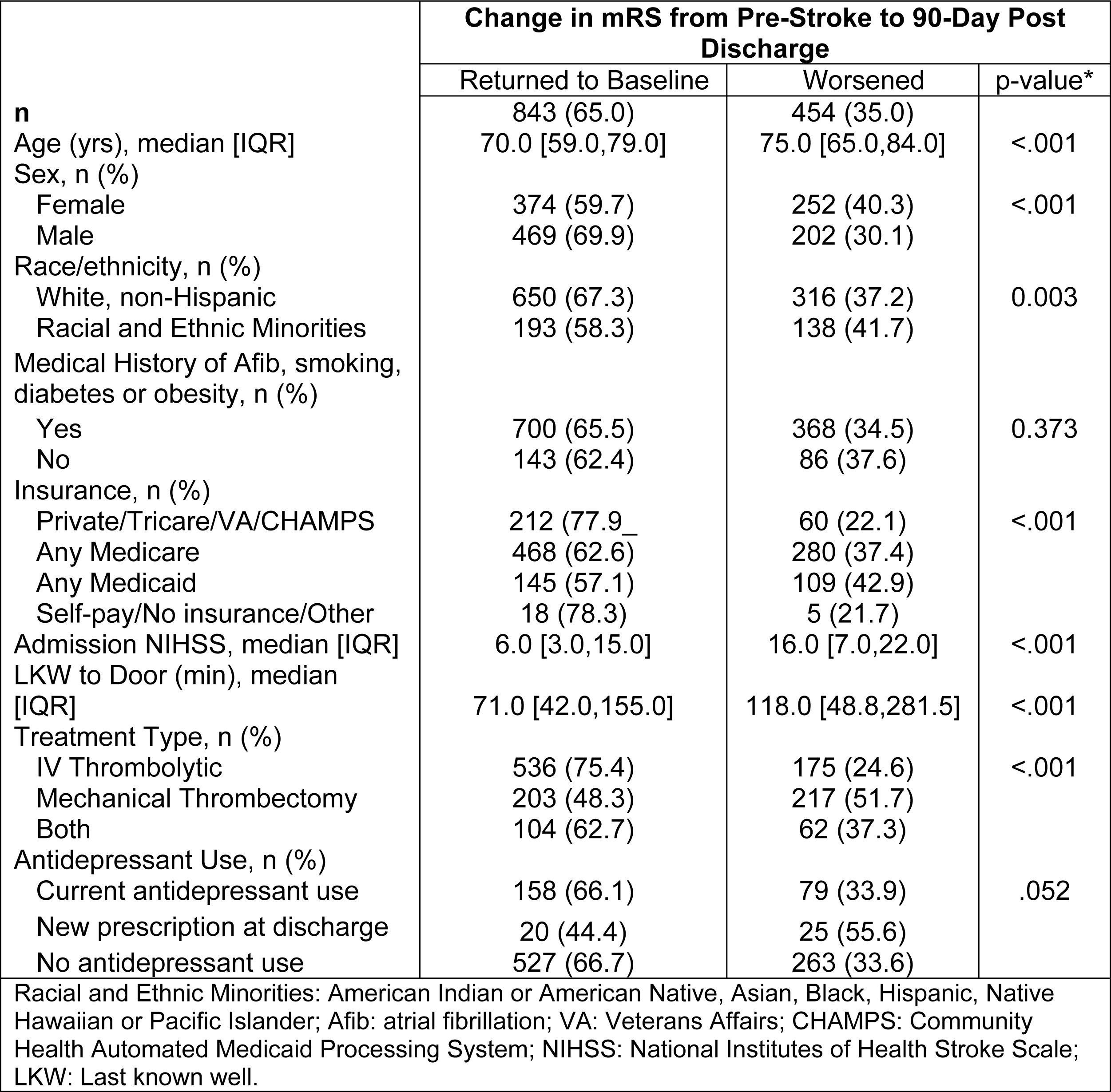
Cohort characteristics by mRS status at 90-days.

**Table 6:** Model of change in mRS (pre-stroke to 90-days post discharge)

| Antidepressant Treatment | Unadjusted Odds Ratio |  |  | Adjusted Odds Ratio* |  |  |
| --- | --- | --- | --- | --- | --- | --- |
|  | OR | 95% CI | p | OR | 95% CI | p |
| No current antidepressant use<br>(Reference=Current antidepressant use) | 0.939 | 0.739-1.20 | 0.611 | 0.955 | 0.638-1.34 | .789 |
| No current antidepressant use | 1.00 | 0.801-2.70 | .987 | 0.989 | 0.763-1.28 | .931 |
| New antidepressant prescription at discharge<br>(Reference=Current antidepressant use) | 0.400 | 0.261-0.614 | <.001 | 0.510 | 0.277-0.938 | .030 |
| mRS: Modified Rankin Scale, CI: Confidence Interval, OR: Odds Ratio |  |  |  |  |  |  |
| * Model adjusted for age, sex, race and ethnicity, NIHSS at admission, time from last know well to arrival, previous medical history of diabetes, AFIB, smoking or obesity, insurance, and stroke treatment type. |  |  |  |  |  |  |

### 3.5 Post-Hoc Analysis of Antidepressant Use and Sex-Specific Interactions

In the post hoc analyses, only change in functional independence at 90-days post-discharge had interactions between antidepressant use and the model covariates. Furthermore, only sex was significant in the two-way interactions. Females with current antidepressant use had a higher predicted probability of returning to baseline compared to females with no antidepressant use (Figure 3a). This finding did not exist for males. Overall, males with no antidepressant use had the highest predicted probability of returning to baseline at 90-days. However, the difference was only significant compared to males who were prescribed an antidepressant for the first time at discharge, the group with the lowest predicted probability of returning to baseline. When comparing females to males with the same antidepressant use, the only significant difference was found for patients with no antidepressant use (Figure 3b).

**Figure 3.**
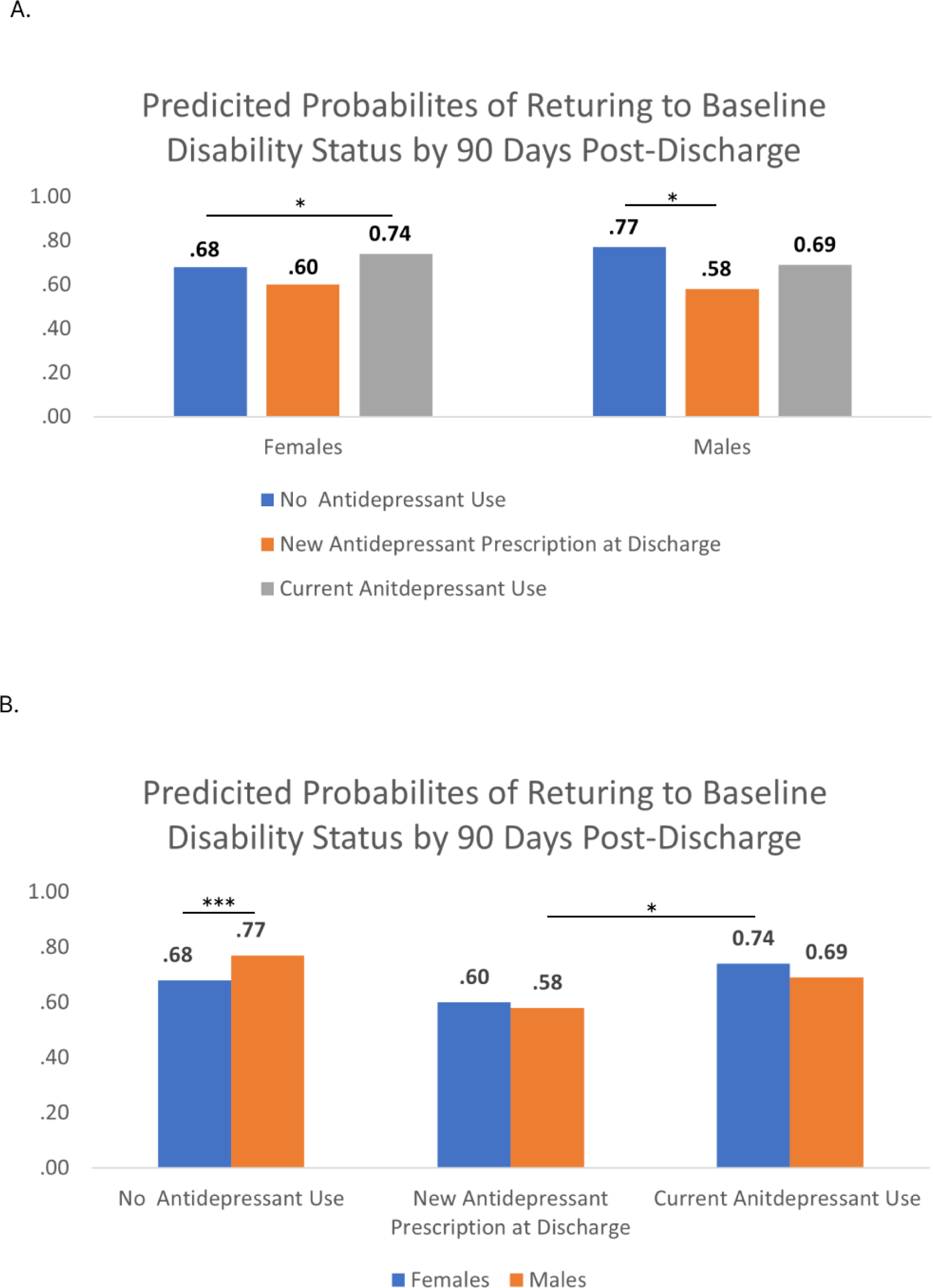
Sex interaction with antidepressant use on functional independence at 90-days post-discharge: Predicted probabilities of returning to baseline functional independence by 90-days post-discharge for females and males by antidepressant use are shown. No antidepressant use: females, n = 339; males, n = 451. New antidepressant prescription at discharge: females, n = 24; males, n = 26. Current antidepressant use: females, n = 157; males, n= 80. **(A)** Compares predicted probabilities of returning to baseline functional independence between different antidepressant groups within the same sex. **(B)** compares the predicted probabilities of returning to baseline functional independence for males and females with the same antidepressant use. (*P ≤ 0.05, ***P ≤ 0.001).

## 4.0 Discussion

We found that patients with current antidepressant use were associated with a decreased likelihood of returning to baseline ambulation and functional independence at discharge than any other group. These results corroborate a previous study that found functional outcomes at discharge improved among patients treated with antidepressants prior to stoke compared to those who received treatment during hospitalization.[10] However, at 90-days post-discharge, we found that patients with no history of antidepressant use or with a previous history of antidepressant use had “caught up” with patients currently taking antidepressants at time of stroke. In contrast, patients who were prescribed a de novo antidepressant at discharge were associated with a ∼57% decreased likelihood of returning to baseline functional independence compared to patients currently using an antidepressant.

The reason for this is unclear. One possible explanation is that those with a new diagnosis of depression may be less adherent to post-stroke therapies, including antidepressants, which could have an impact on overall recovery.[12–15] Another explanation is that the neuroprotective effects of antidepressants are greater if taken pre-stroke compared to post-stroke.[10] Antidepressants are known to have delayed efficacy, with noticeable mood improvement taking 2-4 weeks to emerge.[16] It’s possible that the neuroprotective effects of antidepressants are also delayed. Additionally, evidence from real-world settings show that ∼50-60% of patients diagnosed with depression do not respond to the first antidepressant medication they are prescribed, which may impact stroke recovery. [17,18]

We also found that sex had a significant interaction in our model on the impact of antidepressant use on functional outcomes at 90-days. Among females, those with current antidepressant use had higher predicted probability of returning to baseline compared to those who had never been on an antidepressant. One potential explanation for this is that females treated with antidepressants prior to a stoke are more likely to experience neuroprotective or neurogenerative effects that lead to improved outcomes compared to females who are not.[19–21] A second possible explanation is that women who are treated with antidepressants may have greater resources or access to healthcare than those who are not, which translates to improved recovery.[19–21] We did not have data on socioeconomic status (SES). However, we did include insurance as a proxy for SES and access to care. Neither insurance alone nor the interaction with antidepressant treatment were significant predictors of outcomes.

Among males, those with a first-time prescription at discharge associated with a decreased likelihood of returning to baseline at 90-days compared to those with no antidepressant use. Moreover, this group had the lowest predicted probability of returning to baseline among all the treatment groups by sex. Males who are newly diagnosed with depression may not respond well or adhere to antidepressant treatment, which can affect recovery.[22,23] Although more study is needed to clarify the mechanisms, our study suggests that clinicians should be cognizant of acute ischemic stroke patients’ history with antidepressant use. For those that are newly diagnosed after their stroke and provided with an antidepressant at discharge, close follow-up by a provider may be warranted to ensure that optimal recovery is attained.

Our observational study is limited by several factors. First, we do not know what antidepressant drug the patients in our cohort used or why they were prescribed antidepressants. Therefore, we cannot determine the impact that specific drugs or conditions may have on stroke outcomes. Second, we analyzed race and ethnicity as a dichotomized variable due to small sample sizes of racial and ethnic minority patients. We ran the models with more granular racial and ethnic groups. However, since the groups were small, the model results were not stable given the small cell sizes. Furthermore, inclusion of the expanded variable did not change the trajectory of the independent or dependent variables of interest. As a result, even though our cohort reflected local demographics at the study site, we cannot generalize our findings to all racial and ethnic groups or to regions within the United States with different demographic make-ups. Third, not all treated patients had an assessment of functional independence at 90-days post-discharge. As this was an observational study, whether patients received a 90-day follow-up differed by hospital protocols and depended on stroke providers’ clinical judgement. Therefore, while our two mRS cohorts are large, they do not represent an entirely random sampling of the general stroke population. While the patient characteristics between our total population and the outcome cohorts were similar (Table 1), the results from our mRS outcomes may not be fully generalizable. Fourth, our study does not account for how post-discharge care, such as physical therapy and rehabilitation, influences functional outcomes. However, a 2020 review showed that most clinical trials of motor-rehabilitation after stroke show no significant improvement between experimental and control groups.[24] Finally, we only included thrombolytic or thrombectomy treatment patients in the analyses, so these results are not generalizable to patients who did not receive these stroke treatments. Untreated stroke patients were excluded because they have very different functional outcomes compared to treated stroke patients. Including them would have confounded our ability to determine the effect of antidepressant use on outcomes.

## 5.0 Conclusions

Our study is a large, multi-center investigation of real-world data to examine the association of the timing of antidepressant use with ambulatory and functional outcomes post-stroke. The results support that the use of antidepressants prior to an acute ischemic stroke associates with stroke recovery, but the effect is moderated by sex. In addition, we found that patients prescribed a new antidepressant at discharge had the poorest outcomes at 90-days. Further study is needed to determine the relationship and mechanisms between timing of antidepressant treatment and outcomes.

## Statement of Ethics

The Institutional Review Board of Providence Health and Services approved this study with a waiver of informed consent.

## Conflict of Interest Statement

The authors have no conflicts of interest to declare.

## Funding Sources

The study was funded internally by Providence St Joseph Health.

## Author Contributions

Elizabeth Baraban contributed to the conception of the research idea, study design, conducting and interpreting the analyses, writing and revision the manuscript for intellectual content.

Kyle Still contributed to the writing and revision of the manuscript for intellectual content.

Alexandra Lesko contributed to the study design, data interpretation, and the writing and revision of the manuscript for intellectual content.

Weston Anderson contributed to the interpretation of data, writing the original draft, and revision of the manuscript for intellectual content.

All authors approved of the final version of the manuscript to be published.

## Data Availability Statement

Non-PHI data may be made available from the corresponding author on reasonable request and upon approval by Providence Institutional Review Board and Providence St Joseph Health leadership.

## Notes

### Competing Interest Statement

The authors have declared no competing interest.

